# Inter-scanner reproducibility of brain multifrequency MR elastography

**DOI:** 10.64898/2026.04.13.26350765

**Authors:** Simon Murk, Frederik B. Laun, Stefan Rampp, Martin Vossiek, Jakob Schattenfroh, Jing Guo, Ingolf Sack, Arnd Doerfler, Guillaume Flé

## Abstract

**Background:** Brain magnetic resonance elastography (MRE) is an emerging quantitative neuroimaging technique that provides noninvasive maps of brain tissue viscoelasticity. For multi-center applications, robust cross-site reproducibility across scanner platforms is essential but remains insufficiently characterized.

**Purpose:** To evaluate cross-site reproducibility of brain multifrequency MRE measurements between two MRI scanner platforms using harmonized protocols.

**Study Type:** Prospective cross-site test-retest reproducibility study.

**Study Population:** Sixteen healthy adult volunteers (7 men, 9 women; mean age 32.2 ± 8.0 years).

**Field Strength/Sequence:** 3 T systems (Siemens MAGNETOM Cima.X and MAGNETOM Vida at two sites) with identical brain multifrequency MRE sequences, echo-planar imaging (EPI) readout, and standardized driver configuration.

**Assessment:** Each participant underwent one MRE acquisition at each site. Shear wave speed (SWS) and penetration rate (PR) were quantified in whole brain, white matter, subcortical gray matter, and cortical gray matter regions using atlas-based region-of-interest (ROI) analysis in MNI152 space.

**Statistical Tests:** Absolute relative difference (ARD), reproducibility coefficient (RDC), coefficient of variation (CV), intraclass correlation coefficient (ICC), and Bland-Altman plots were calculated to determine cross-site reproducibility.

**Results:** Cross-site reproducibility was robust for major brain regions, with region-averaged ARD values for SWS ranging from 1.38 % to 3.43 % and for PR from 3.20 % to 7.25 % across tissues. RDCs for SWS ranged from 0.02 m.s^-1^ to 0.07 m.s^-1^, and for PR from 0.03 m.s^-1^ to 0.08 m.s^-1^. Coefficients of variation for SWS ranged from 0.82 % to 1.93 %, and for PR from 2.21 % to 4.09 %. ICC values for SWS ranged from 0.66 to 0.84 and for PR from 0.67 to 0.88. Bland-Altman analysis showed minimal systematic bias and tight limits of agreement.

**Conclusion:** Brain multifrequency MRE demonstrates robust reproducibility across distinct 3 T platforms when using harmonized acquisition and reconstruction. These results support the use of brain MRE as a quantitative biomarker and provide benchmark reproducibility metrics for future research.

## 1 Introduction

Magnetic resonance elastography (MRE) is a quantitative magnetic resonance imaging (MRI) method that noninvasively determines the mechanical properties of soft tissue by analyzing the propagation of externally induced mechanical waves imaged with specialized phase-contrast magnetic resonance (MR) techniques. Over the past decade, advances in MRE hardware, MR pulse sequences, and inversion algorithms have enabled the generation of high-resolution mechanical property maps, notably in the brain, which have facilitated the assessment of aging-related brain tissue alterations [1, 2, 3], Alzheimer’s disease [4, 5, 6, 7], multiple sclerosis [8], Parkinson’s disease [9, 10] and neuro-oncology [11, 12, 13]. While MRE is intrinsically sensitive to mechanical contrasts and disease-specific alterations, the resulting property maps are also influenced by sequence parameters [14, 15, 16, 17], vibration frequency [18, 19, 20, 21], and reconstruction methods [22, 23, 24, 25]. These dependencies complicate cross-study comparisons and the evaluation of MRE’s stability for clinical integration, as measurements often rely on heterogeneous scanner hardware, MRE parameters, and software environments.

Repeatability studies using fixed and varied protocols have contributed to demonstrating the robustness of MRE in healthy volunteer cohorts measured multiple times at a single MR device [24, 26, 27]. These studies showed good-to-excellent within-session repeatability in brain MRE. In addition to repeatability assessments, atlases of brain viscoelastic properties mapped in standard space, e.g., MNI152 [28], and derived from larger participant cohorts, measured across single sites, provide a valuable framework for consistency analysis of viscoelasticity values in specific tissue compartments [29, 30]. However, the reproducibility of brain MRE has not been assessed in a multi-site setting using the same participants. This is especially critical for clinical translation, where multi-site trials may involve data from different MR scanners.

Our study addresses this need and systematically assesses cross-site reproducibility of brain MRE in a cohort of healthy adults who underwent scans on two distinct 3 T MRI platforms with identical multifrequency spin-echo EPI-based MRE sequences [31], a standardized mechanical driver configuration, and a multifrequency wavenumber-based inversion pipeline (*k*-MDEV) [32]. We quantify shear wave speed (SWS), which indicates tissue stiffness, and penetration rate (PR), which quantifies the attenuation of shear wave amplitude due to viscous and geometric damping, within atlas-based tissue compartments derived from a multi-step processing workflow, which includes correction procedures and multi-stage registration. This work uses absolute relative differences (ARD), reproducibility coefficients (RDC), coefficients of variation (CV), intraclass correlation coefficients (ICC), and Bland-Altman plots to establish quantitative benchmarks for cross-site reproducibility in brain MRE measurements. These benchmarks provide a foundation for future multi-site studies.

## 2 Material and Methods

### 2.1 Study design and participants

Sixteen healthy adults participated in this prospective cross-site reproducibility study (7 men, 9 women; mean age 32.2 ± 8.0 years; range 23–58 years). The study protocol received approval from the institutional ethics committee of the Friedrich Alexander Universität Erlangen Nürnberg (application 23-389-Bm). All participants provided written informed consent prior to study participation. Inclusion criteria comprised age ≥18 years, absence of neurological or cardiovascular disease, no regular medication, and no contraindications to MRI. Exclusion criteria comprised prior major head injury or neurosurgical intervention, clinically relevant alcohol abuse, pregnancy, and the presence of any MRI-incompatible implanted device.

### 2.2 Experimental setup

MRE acquisitions were performed on two 3 T MRI systems (Siemens MAGNETOM Cima.X, *Scan A*; Siemens MAGNETOM Vida, *Scan B*) equipped with identical 64 channel head coils. The scanners were located at two distinct sites of the University Hospital Erlangen (Germany). Figure 1 illustrates the experimental setup. Mechanical vibrations were generated using an active pneumatic driver powered by compressed air (VibroR42, THEA Devices GmbH, Germany) and were transmitted via plastic hoses to two pillow-like passive transducers operated in phase opposition and positioned over the occipital area of the scalp at approximately the 5 and 7 o’clock positions relative to the head. Head position and transducer placement followed a standardized protocol and were maintained as consistently as possible across scans and sites. All participants underwent imaging first at *Site A*, followed by *Site B*. For most participants, both acquisitions were conducted on the same day with a 2 to 4-hour interval between scans. During this interval, participants remained unsupervised. In two participants, same-day acquisition was not feasible. In these cases, the interval between scans was 7 and 176 days. No distinct behavior of these subjects compared to the remaining data was observed.

**Figure 1:**
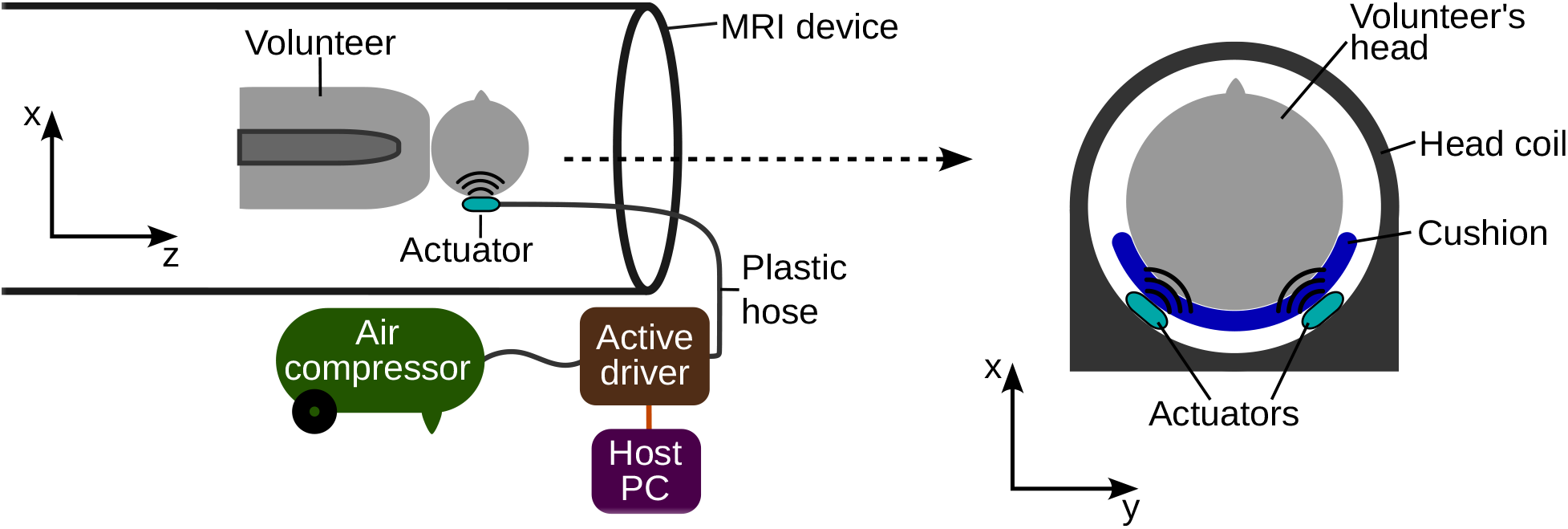
Experimental setup.

### 2.3 Image acquisition

At both sites, identical versions of a multifrequency MRE sequence incorporating motion-encoding gradients (MEG) with first moment nulling and echo-planar readout were used to acquire three-dimensional brain tissue displacement data [31]. Mechanical vibrations frequencies were 20, 25, 30, and 35 Hz, as in [24]. Vibration amplitude was controlled with corresponding pneumatic pressure levels of 175, 225, 250, and 350 mbar. External vibrations were synchronized with the motion-encoding gradients (MEGs) of the MRE sequence by calibrating the digital clocks of wave generator to that of the MRI scanner. Otherwise no timing synchronization was performed by the wave generator while the MRE sequence timing was adapted relative to the external wave phase to sample eight evenly spaced timepoints over one vibration cycle for each frequency [31]. Motion encoding was performed along the three orthogonal spatial directions (*x, y*, and *z*). In total, 96 MRE head volumes were acquired, corresponding to the four vibration frequencies, eight timepoints, and three MEG directions. MRE acquisition parameters included a repetition time (TR) of 6540 ms, echo time (TE) of 70 ms, an acquisition matrix of 116 × 116, 60 slices, an isotropic voxel size of 2.0× 2.0× 2.0 mm^3^, and a phase-encoding (PE) direction along the anteroposterior axis. Each imaging session included the measurement of EPI volume pairs acquired with reversed PE directions, i.e., anteroposterior and posteroanterior, for subsequent correction of geometry distortion induced by material heterogeneous susceptibility distributions and long EPI readouts [33]. Apart from the PE direction, all imaging parameters remained identical to those of the MRE sequence. For distortion correction, a total of six EPI volumes were acquired, corresponding to two PE and three MEG directions. For anatomical reference, T1-weighted structural images were acquired using a magnetization-prepared rapid gradient echo sequence (MPRAGE). The sequence parameters of the T1-weighted sequence included a TR of 2200 ms, TE of 2.46 ms, an acquisition matrix of 256 × 256, 208 slices, and an isotropic voxel size of 0.8 × 0.8 × 0.8 mm^3^. Sequence geometry, vibration driver configuration, and relevant system parameters were documented and harmonized across scanners to ensure protocol consistency between sites.

### 2.4 Data processing and MRE inversion

Both MRE magnitude and phase data were processed using a multi-step post-processing pipeline to obtain quantitative maps of SWS and PR. The processing workflow was implemented in MATLAB (version R2024b, Natick, Massachusetts, US) and is illustrated in Fig. 2. First, wrapped phase images were unwrapped using a Laplacian-based phase unwrapping algorithm (*Step 1*). This method was originally described by Dittmann *et al*. and is provided in the appendix of reference [31]. Second, subject motion was corrected using an in-plane two-dimensional motion correction procedure implemented in the statistical parametric mapping toolbox (SPM,,version 25.01.02, University College London, UK; *Step 2*) [34]. Here, the first head volume was used as a reference for the remaining 95 volumes. Third, geometric distortions were corrected using TOPUP from the FMRIB software library (FSL, version 6.0.7.17, Oxford University, UK; *Step 3*) [35]. Fourth, brain masking was performed using the brain extraction module (BET) implemented in FSL to remove non-brain structures and restrict subsequent analysis to intracranial tissue (*Step 4*). Fifth, wave motion at the applied driver frequency was extracted from the time-resolved phase data using the fast Fourier transformation (tFFT; *Step 5*). Sixth, interslice phase correction was performed to compensate for phase offsets between slices and ensure consistent wave fields throughout the three-dimensional volume (*Step 6*). Finally, mechanical property reconstruction was performed using the wavenumber-based multifrequency dual-elasto-visco method (*k*-MDEV), which is publicly available online at https://bioqic.charite.de (*Step 7*) [25, 32]. This inversion procedure generated quantitative maps of SWS and PR by estimating the complex-valued wavenumber *k*^∗^ = *k*^*′*^ + *ik*^*′′*^ from a plane-wave decomposition of the measured wave field at a single vibration frequency. Real and imaginary parts of the wavenumber are given by [24]:

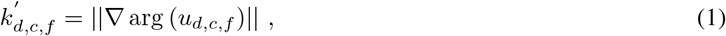

and

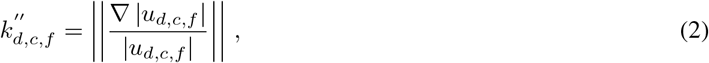

where indices *d, c*, and *f* refer to the plane wave direction of propagation, the displacement vector component, and the vibration frequency, respectively.

**Figure 2:**
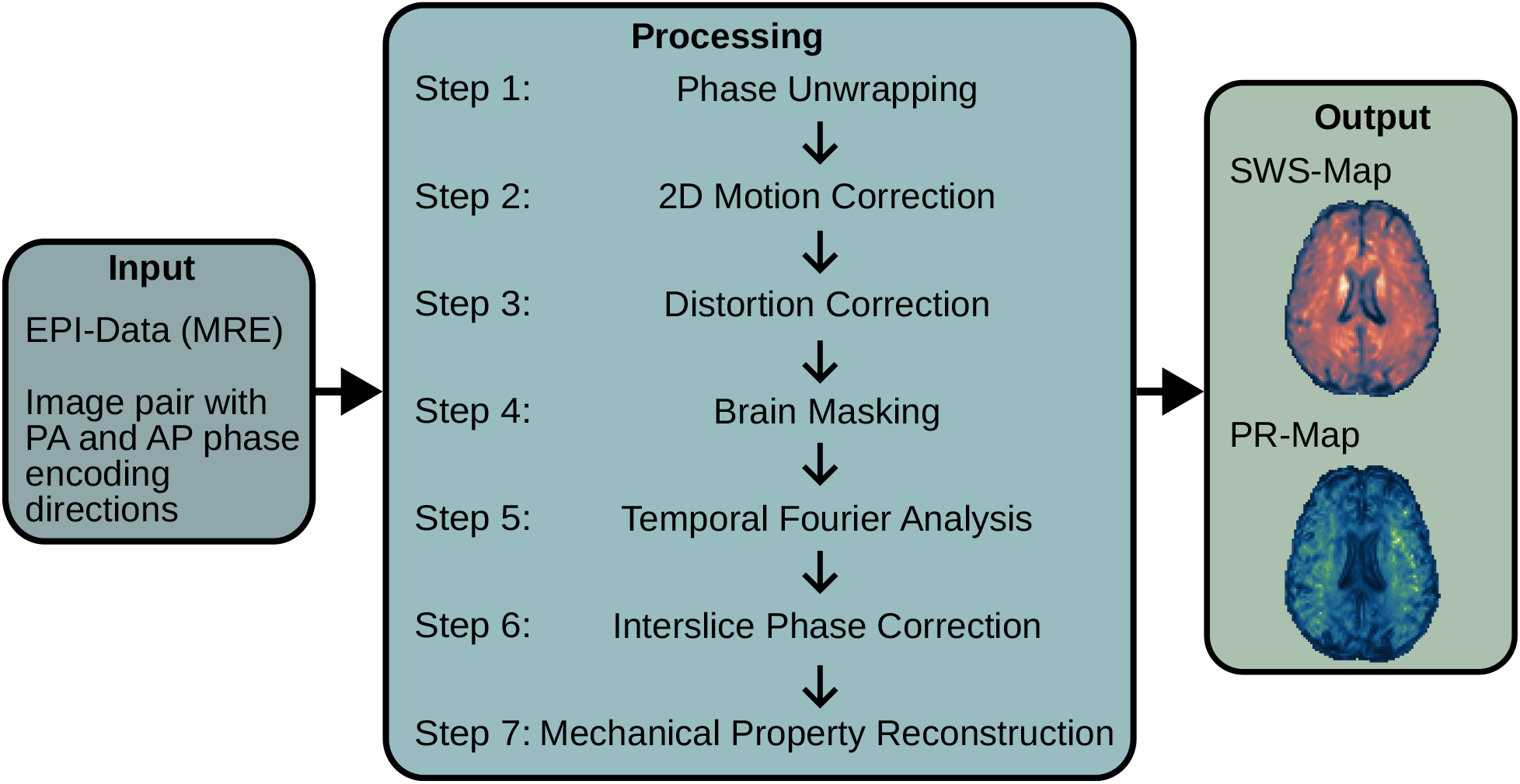
Data processing. Abbreviations: SWS, shear wave speed; PR, penetration rate.

Frequency-resolved SWS_*f*_ and PR_*f*_ were calculated by [24]:

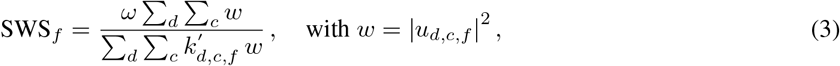

and

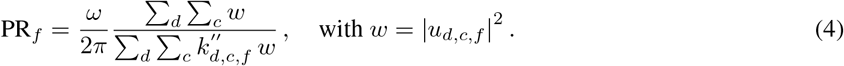

Final SWS and PR distributions were obtained by averaging the frequency-resolved SWS_*f*_ and PR_*f*_ volumes. The resulting SWS and PR maps were initially reconstructed in EPI native space.

*Abbreviations: SWS, shear wave speed; PR, penetration rate*.

### 2.5 Image registration and spatial normalization

For the regional comparisons between scans, mechanical property maps were spatially normalized to standard space. Image registration was implemented in a multi-step MATLAB pipeline that used FSL tools for all transformation procedures and is illustrated in Fig. 3. First (*Registration 1*), a linear within-subject registration aligned the skull-stripped (denoted by *) MPRAGE image of *Scan B* to the corresponding image of *Scan A* using FSL FLIRT with six degrees of freedom (T1w.B* →T1w.A*). The resulting pre-aligned (denoted by ‘) image (T1w.B’), served as the reference for subsequent processing steps of *Scan B*. Second (*Registration 2A/B*), for each session separately, the skull-stripped magnitude EPI image was linearly registered to the corresponding skull-stripped MPRAGE image using FLIRT with six degrees of freedom (EPImag.A*/B*→ T1w.A*/B’*). Third (*Registration 3A/B, Step 1*), each skull-stripped MPRAGE image (T1w.A* and T1w.B’*) was linearly registered to the MNI152 T1-weighted 2 mm brain template using 12 degrees of freedom to account for global scaling and shear (T1w.A*/B’* →MNI152*). Fourth (*Registration 3A/B, Step 2*), a non-linear warp from non-skull-stripped MPRAGE image to the MNI152 T1-weighted 2 mm template was estimated with FSL FNIRT to refine anatomical correspondence (T1w.A/B →MNI152). The affine transformation matrix obtained in *Step 1* served as the initial transformation for the non-linear registration. Finally, the linear transformations and the non-linear warp fields were concatenated into a composite transformation using FSL APPLYWARP and applied to the SWS and PR maps of each session (SWS.A/B →MNI152; PR.A/B →MNI152). In this step, spline interpolation was used to minimize resampling artifacts. This procedure yielded mechanical property maps normalized to MNI152 standard space with an isotropic voxel size of 2 mm. All normalized images underwent visual quality control with particular attention to residual misregistration. The workflow depicted in Fig. 3 is to be understood such that the images or transformations specified along each arrow are propagated to and used as inputs for the subsequent processing step.

**Figure 3:**
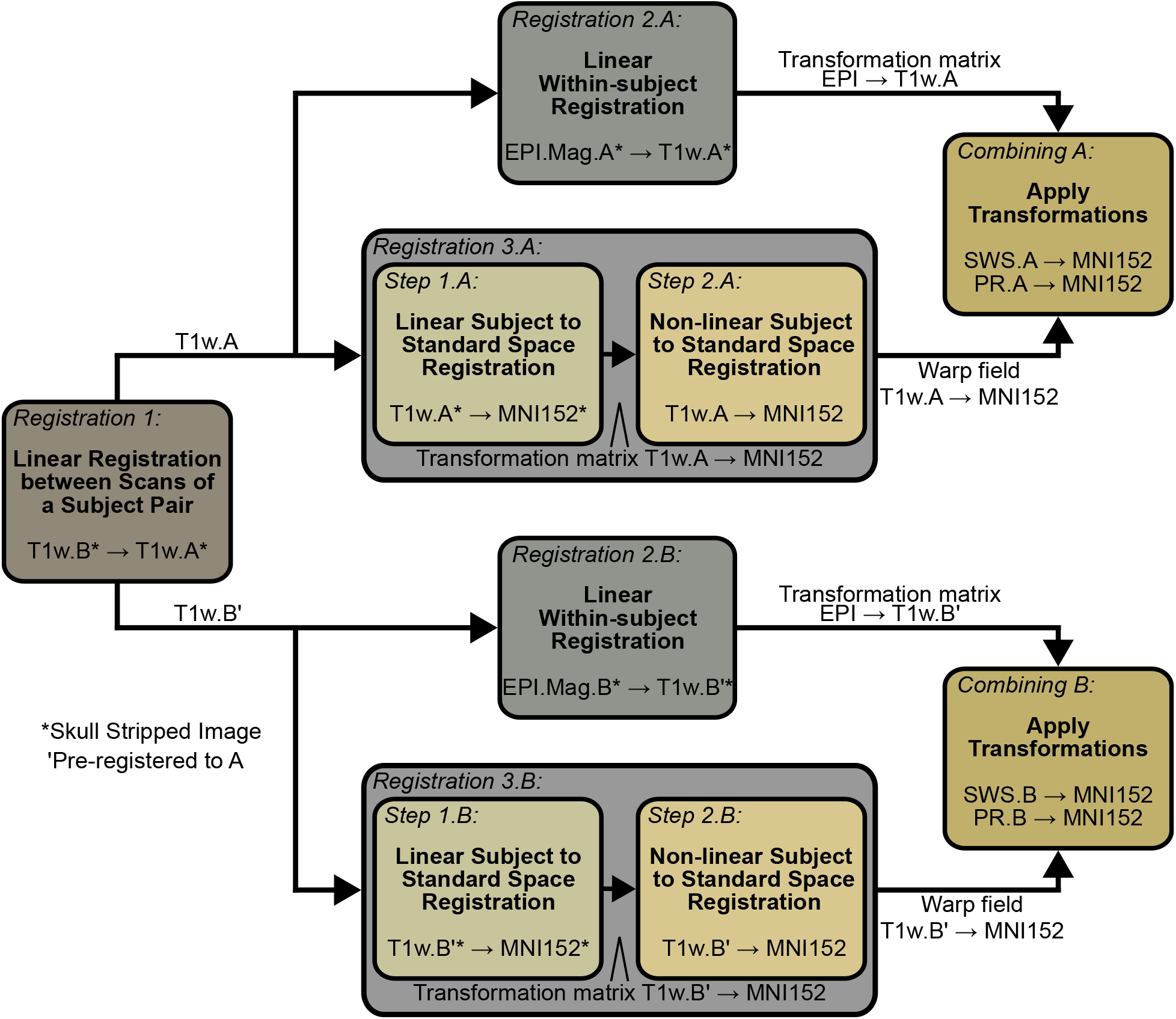
Registration process.

### 2.6 Region-of-interest analysis

Region-of-interest analysis (ROI) was performed in MNI152 standard space and followed a compartment-based approach similar to that described by Hiscox *et al*. (2020) [30]. Tissue compartments masks were derived from atlas-based probability maps. ROIs included a whole brain ROI based on the MNI152 brain mask, a white matter ROI derived from the MNI white matter map (threshold ≥ 0.75), a subcortical gray matter ROI derived from the Harvard-Oxford subcortical atlas (threshold ≥ 0.75), and a cortical gray matter ROI derived from the Harvard-Oxford cortical atlas (threshold ≥ 0.65). Mean SWS and PR values were extracted from each ROI for every subject and scan. Representative slices of the resulting ROI masks are illustrated in Fig. 4.

**Figure 4:**
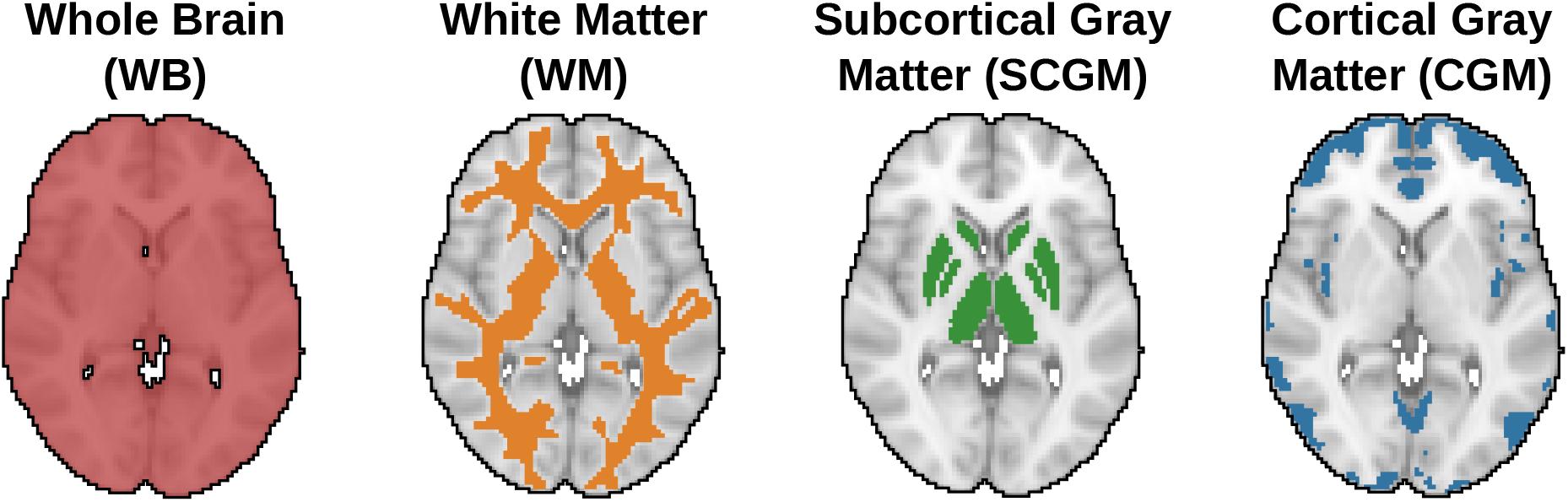
Color coded visualization of the four brain regions analyzed: whole brain, white matter, subcortical gray matter, and cortical gray matter.

### 2.7 Statistical analysis

Multiple quantitative metrics were calculated to comprehensively assess reproducibility. Absolute relative differences (ARD) were used to quantify pairwise deviations between repeated measurements and are given by:

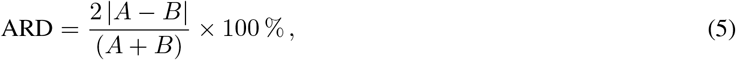

where A and B represent the measured values from *Scan A* and *B*, respectively. ARD values were calculated voxel-wise and spatially averaged in the individual brain regions shown in Fig. 4. Similarly, symmetric relative differences (SRD) were calculated to visualize variation trends between the two scans and are given by:

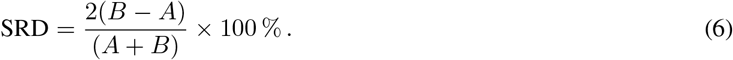

The reproducibility coefficient (RDC), which represents the 95 % confidence limit for expected measurement variation in original units, was calculated as:

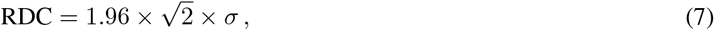

where *σ* denotes the within-subject standard deviation. Relative variability was further assessed using the coefficient of variation (CV) [36]:

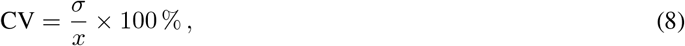

where *x* is the average of all measurements. In addition, the intraclass correlation coefficient (ICC) was computed as [37]:

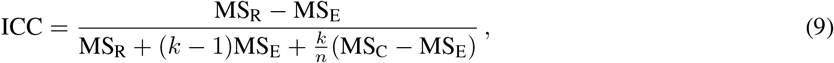

*Abbreviations: SWS, shear wave speed; PR, penetration rate; ROI, region of interest; ARD, absolute relative difference; SD, standard deviation; WB, whole brain; WM, white matter; SCGM, subcortical gray matter; CGM, cortical gray matter*.

where MS_R_ represent the mean square between subjects, MS_C_ the mean square between scanners, MS_E_ the residual (error) mean square, *k* the number of scanners, and *n* the number of subjects. ICC values of 0.50-0.75 were interpreted as moderate agreement across all data sets, whereas values of 0.75-0.90 indicated good agreement [37]. Bland-Altman plots were generated to evaluate systematic bias, 95 % limits of agreement, and visually assess the agreement between the repeated measurements [38, 39]. All statistical analyses were performed using MATLAB (version R2025b) with the Statistics and Machine Learning Toolbox (version 25.1) and a custom ICC MATLAB function (version 1.3.1.0) [40]. Mean ARD, RDC, CV, and ICC were reported for each ROI, with 95 % confidence intervals (CIs) for RDC and ICC, and standard deviations (SDs) for ARD.

## 3 Results

### 3.1 Absolute relative difference

Across mechanical parameters and brain regions (whole brain, white matter, subcortical gray matter, and cortical gray matter), mean absolute SWS values in *Scan A* were 1.07 ± 0.03 (WB), 1.25± 0.02 (WM), 1.33 ± 0.07 (SCGM), and 0.99 ± 0.03 m.s^-1^ (CGM) and in *Scan B* 1.08 ± 0.03 (WB), 1.26 ± 0.02 (WM), 1,37 ± 0.06 (SCGM), and 1.00 ± 0.03 m.s^-1^ (CGM). Mean absolute PR values in *Scan A* were 0.64 ±0.04 (WB), 0.79 ± 0.04 (WM), 0.66 ± 0.06 (SCGM), and 0.57 ± 0.04 m.s^-1^ (CGM) and in *Scan B* 0.65± 0.04 (WB), 0.81 ± 0.03 (WM), 0.70 ± 0.06 (SCGM), and 0.58 ± 0.04 m.s^-1^ (CGM). Mean ARD values ranged from 1.38 % (SWS in white matter) to 7.25 % (PR in subcortical gray matter). Cortical gray matter demonstrated the smallest ARD for PR (3.20 ± 2.22 %). In contrast, subcortical gray matter showed the highest ARD values for both parameters, with 3.43 ± 2.25 % for SWS and 7.25 ± 3.84 % for PR. Across all regions, ARD values were consistently lower for SWS than for PR. A summary of ARD values for all analyzed brain regions is presented in Table 1.

**Table 1:** Mean and SD of ARD values for SWS and PR across ROIs.

| ROI | SWS |  | PR |  |
| --- | --- | --- | --- | --- |
|  | Mean ARD [%] | SD of ARD [%] | Mean ARD [%] | SD of ARD [%] |
| WB | 1.43 | 0.71 | 3.69 | 1.83 |
| WM | 1.38 | 0.84 | 3.44 | 1.83 |
| SCGM | 3.43 | 2.25 | 7.25 | 3.84 |
| CGM | 1.52 | 0.91 | 3.20 | 2.22 |

*Abbreviations: SWS, shear wave speed; PR, penetration rate; ROI, region of interest; ARD, absolute relative difference; SD, standard deviation; WB, whole brain; WM, white matter; SCGM, subcortical gray matter; CGM, cortical gray matter.*

Figure 5 illustrates the spatial agreement between *Scan A* and *Scan B* across the brain. The subject with the lowest whole-brain ARD (0.27 %) showed uniformly low SRD values across cortical and deep gray matter regions. In contrast, the subject with the highest whole-brain ARD (3.00 %) demonstrated a more heterogeneous spatial pattern with locally elevated differences in periventricular areas and deep gray matter structures.

**Figure 5:**
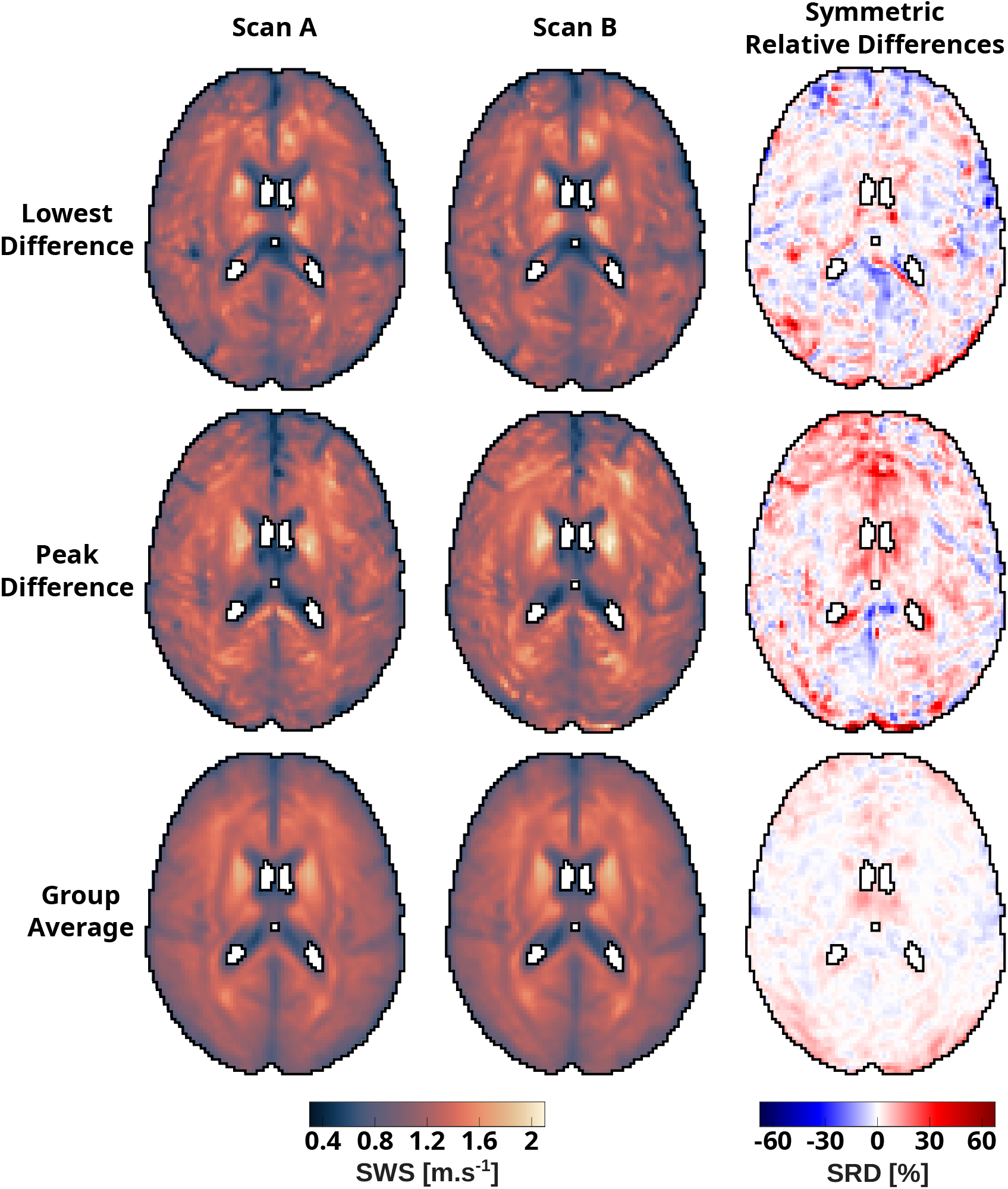
SWS (left and middle) and SRD maps (right) for the subject pairs with the smallest (0.27 %) and largest (3.00 %) whole brain averaged ARD value. The bottom row shows SWS and SRD average maps over all subjects. Abbreviations: SWS, shear wave speed; PR, penetration rate; SRD, symmetric relative difference.

### 3.2 Analysis of mechanical property distributions in the brain

Figure 6 shows the distribution of region-averaged SWS and PR values across subjects for each site and ROI using violin plots. The size of the violin plots is proportional to the number of data points, the horizontal solid line indicates the mean value of the material property, and the dashed lines indicate the limits of the Interquartile Range. While not totally symmetrical, the violin plots showed similar distributions for scans at both sites in the four analyzed brain regions. For SWS, the mean value was marginally higher in *Scan B* by 0.01, 0.01, 0.04, and 0.01 m.s^-1^ for the whole brain, white matter, subcortical gray matter, and cortical gray matter, respectively. For PR, the mean value was also higher in *Scan B* by 0.01, 0.02, 0.04, and 0.01 m.s^-1^ for the whole brain, white matter, subcortical gray matter, and cortical gray matter, respectively.

**Figure 6:**
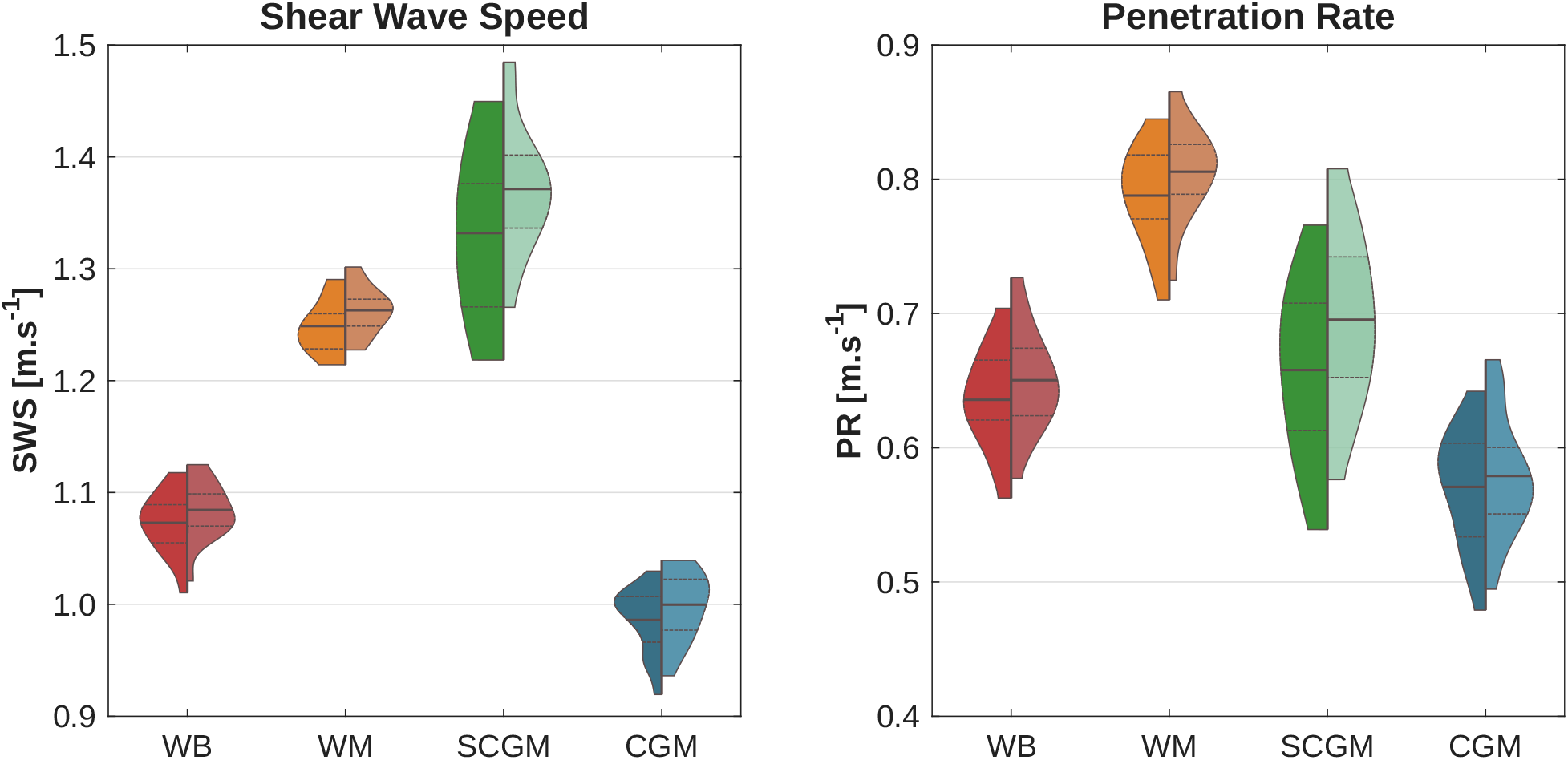
Split violin plots of region-averaged SWS and PR values of *Scan A* (left-hand half) and *Scan B* (right-hand half) for four brain regions. Abbreviations: SWS, shear wave speed; PR, penetration rate; WB, whole brain; WM, white matter; SCGM, subcortical gray matter; CGM, cortical gray matter.

### 3.3 Bland-Altman analysis

The Bland-Altman plots shown in Fig. 7 illustrate the agreement between *Scan A* and *Scan B* for SWS (top row) and PR (bottom row) across all ROIs. For SWS, mean differences remained close to zero in all regions. Slight positive offsets of 0.01 m.s^-1^ appeared in the whole brain, white matter, and cortical gray matter regions, whereas subcortical gray matter showed a small positive offset of 0.04 m.s^-1^. This pattern indicates marginally higher SWS values at *Scan B* in most regions and slightly higher values at *Scan A* in subcortical gray matter. Limits of agreement were narrowest in cortical gray matter (approx. ± 0.02 m.s^-1^) and widest in subcortical gray matter (approx. ±0.07 m.s^-1^). For PR, Bland-Altman plots showed broader limits of agreement and higher scatter, particularly in subcortical gray matter, while mean biases remained small across all ROIs.

**Figure 7:**
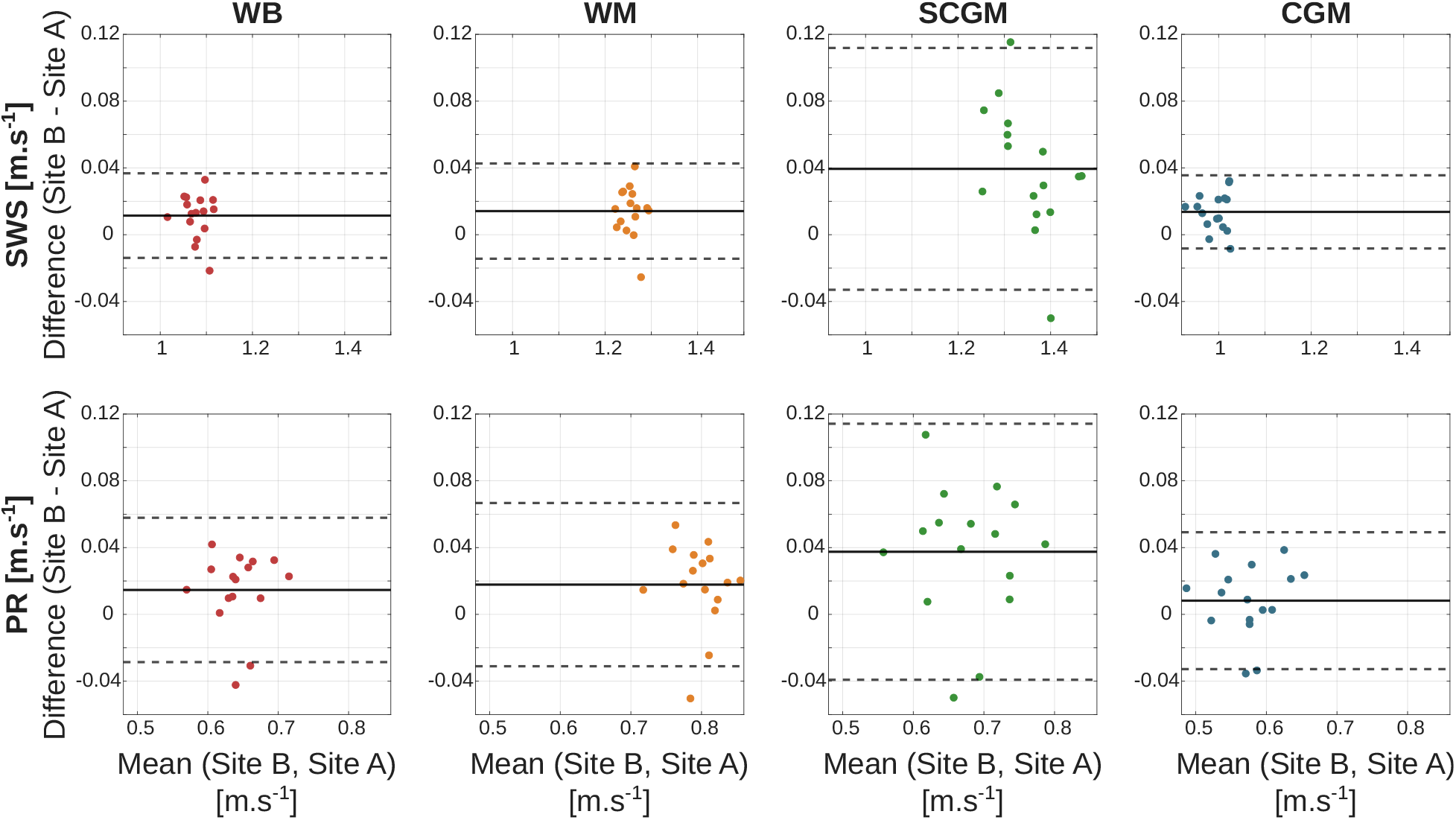
Bland-Altman plots of the SWS and PR obtained in *Scan A* vs. *Scan B* across four brain regions. Top row shows SWS and bottom row shows PR. Mean differences are visualized as continuous thick lines, whereas the limits of agreement are shown as dotted lines. Abbreviations: SWS, shear wave speed; PR, penetration rate; WB, whole brain; WM, white matter; SCGM, subcortical gray matter; CGM, cortical gray matter.

### 3.4 Quantitative reproducibility metrics

The reproducibility metrics for SWS and PR across the four brain regions are presented in Table 2. Reproducibility coefficients for SWS ranged from 0.02 m.s^-1^ in the cortical gray matter ROI to 0.07 m.s^-1^ in subcortical gray matter, while coefficients of variation remain below 2.00 % across all ROIs. For PR, reproducibility coefficients ranged from 0.03 m.s^-1^ in the whole brain ROI to 0.08 m.s^-1^ in subcortical gray matter. Corresponding coefficients of variation ranged from 2.21 % in white matter to 4.09 % in subcortical gray matter. ICCs for SWS ranged from 0.66 in white matter to 0.84 in cortical gray matter. For PR, ICC values ranged from 0.67 in white matter to 0.88 in cortical gray matter.

**Table 2:** Reproducibility metrics for SWS and PR across ROIs.

| ROI | SWS |  |  | PR |  |  |
| --- | --- | --- | --- | --- | --- | --- |
|  | RDC [m.s<sup>-1</sup>]<br>(95% CI) | CV [%]<br>(95% CI) | ICC | RDC [m.s<sup>-1</sup>]<br>(95% CI) | CV [%]<br>(95% CI) | ICC |
| WB | 0.03<br>(0.02–0.04) | 0.85 | 0.81<br>(0.31–0.94) | 0.03<br>(0.02–0.05) | 2.43 | 0.77<br>(0.37–0.92) |
| WM | 0.03<br>(0.02–0.04) | 0.82 | 0.66<br>(0.06–0.89) | 0.05<br>(0.04–0.08) | 2.21 | 0.67<br>(0.20–0.88) |
| SCGM | 0.07<br>(0.05–0.11) | 1.93 | 0.72<br>(0.07–0.91) | 0.08<br>(0.06–0.12) | 4.09 | 0.69<br>(0.10–0.90) |
| CGM | 0.02<br>(0.02–0.03) | 0.80 | 0.84<br>(0.13–0.96) | 0.04<br>(0.03–0.06) | 2.57 | 0.88<br>(0.69–0.96) |

*Abbreviations: SWS, shear wave speed; PR, penetration rate; ROI, region of interest; RDC, repeatability coefficient; CV, coefficient of variation; ICC, intraclass correlation coefficient; CI, confidence interval; WB, whole brain; WM, white matter; SCGM, subcortical gray matter; CGM, cortical gray matter.*

## 4 Discussion

This study evaluated the cross-site reproducibility of multifrequency brain MRE across two 3 T MRI platforms under harmonized acquisition and processing conditions. The presented results demonstrate that brain MRE provides robust measurements of brain viscoelasticity across scanners. Whole brain measurements showed excellent SWS reproducibility with a mean ARD of 1.43 %, RDC of 0.03 m.s^-1^, CV of 0.85 %, and ICC of 0.81, and good PR reproducibility with a mean ARD of 3.69 %, RDC of 0.03 m.s^-1^, CV of 2.43 %, and ICC of 0.77. However, tissue-dependent reproducibility patterns were also observed. Both white matter and cortical gray matter showed lower variability and higher reproducibility (CV_WM, CGM_ *<* 1 %, RDC_WM_ = 0.03 m.s^-1^, and RDC_CGM_ = 0.02 m.s^-1^), whereas subcortical gray matter showed slightly higher variability and lower reproducibility (CV_SCGM_ = 1.93 % and RDC_SCGM_ = 0.08 m.s^-1^). These results suggest that stiffness estimates derived from shear wave speed are more consistent in white matter and cortical gray matter than in subcortical gray matter. This discrepancy may stem from the smaller size and higher stiffness of SCGM, which lead to longer shear wavelengths. These longer wavelengths are inherently more difficult to measure accurately, especially in small areas, and reduce the reliability of stiffness estimates in SCGM. Additionally, the deeper location of SCGM leads to stronger wave attenuation, which reduces the signal quality of measured displacements and thus compromises the reliability of stiffness estimates in this region. Overall, ICC values showed lower agreement compared to the other metrics, likely because ICC is sensitive to between-subject variability and may not entirely account for measurement consistency between the two sites.

Across all regions, SWS demonstrated lower CV and RDC values than PR. Whole-brain PR measurements showed an ICC of 0.77, a CV of 2.43 %, and an RDC of 0.03 m.s^-1^. The difference was most pronounced in subcortical gray matter, where PR reached a CV of 4.09 % and an RDC of 0.08 m.s^-1^ despite a high ICC of 0.69. PR therefore exhibited greater dispersion across scanners than SWS. This difference is likely influenced by the physical interpretation of both parameters. SWS primarily reflects shear wave propagation velocity and therefore represents a stiffness-dominated metric. PR describes attenuation-related properties of the wave field and therefore shows stronger sensitivity to signalto-noise characteristics, regions of low wave amplitudes, and inversion regularization. These factors may contribute to measurement variability across scanners because of variability in exact head positioning in the head coil and contact with the mechanical actuator which cannot be 100 % identical as we are dealing with in vivo human measurements in clinical settings.

The magnitude of cross-site variability must be interpreted in relation to known disease-related effect sizes. Absolute relative differences, calculated using equation 5 from published brain MRE study data, report global stiffness reductions of approximately 8 % to 20 % in neurodegenerative diseases [4, 5, 7, 8, 9, 10, 6] and regional alterations of up to 30 % in brain tumors [12]. In the present study, SWS variability remained below 2 % CV across all major tissue compartments, and mean ARD values remained below 3.5 %. These reproducibility limits are therefore substantially smaller than typical pathological effect sizes. This separation between measurement variability and biological signal indicates that clinically relevant biomechanical alterations are unlikely to be obscured by cross-scanner variability when harmonized protocols are applied.

To date, published repeatability investigations in healthy volunteers have evaluated single-frequency brain MRE on a single scanner platform. These studies reported CV values between 2 % and 6 % together with good repeatability [21, 27, 26]. Multifrequency brain MRE also demonstrated excellent shortand long-term repeatability within the same system [24]. Evidence for in vivo multicenter reproducibility remains, however, limited. A recent phantom study by Ozkaya *et al*. (2023) provided initial multicenter validation under heterogeneous technical conditions. That study included scanners from multiple vendors, two field strengths (1.5 T and 3 T), and different acquisition sequences such as 2D GRE and 2D SE-EPI. Reported stiffness CV values ranged from approximately 5 % to 7 % [14].

The present study extends these observations by demonstrating in vivo cross-scanner reproducibility of a multifrequency MRE protocol under harmonized conditions. Acquisition parameters, driver configuration, reconstruction pipeline, and atlas-based analysis were predefined and matched across scanners prior to the study. The use of scanners from the same vendor enabled strict protocol harmonization and minimized confounding variability related to sequence implementation and reconstruction algorithms. The reported reproducibility metrics therefore represent a controlled benchmark under optimized technical conditions and showed that SWS reproducibility in major tissue compartments approached values reported in several single-site repeatability studies.

Several practical implications arise from these findings. White matter and cortical gray matter are well established reference regions that may serve for quality control procedures or normalization strategies in multi-site studies [2, 27, 30]. Subcortical gray matter metrics remain reproducible but exhibit higher CV and RDC values. Studies that target small biomechanical effect sizes in these regions should therefore interpret measurements with caution. Future work should extend the present harmonization framework to additional vendors, field strengths, acquisition protocols, and viscoelasticity image reconstruction methods to help establish universally applicable reproducibility benchmarks for brain MRE.

## 5 Limitations

This study has several limitations that affect the generalizability of its findings. First, the cohort included only 16 healthy adults within a narrow age range. This limits the transferability of the reported reproducibility metrics to older populations or to patients with neurological disease. According to Koo and Li, at least 30 heterogeneous subjects should be included to obtain stable ICC estimates [37]. Nonetheless, studies involving smaller cohorts have been reported in MRE. For example, brain MRE repeatability studies by Svensson *et al*. (2021) and Herthum *et al*. (2022) reported ICC-based repeatability metrics with sample sizes of 15 and 11 subjects, respectively [24, 27]. Similarly, test-retest studies in pathological brain MRE by Friismose *et al*. (2025) and in adrenal MRE by Webster *et al*. (2025) reported reproducibility metrics in cohorts well below 30 subjects [12, 41]. Nevertheless, larger, and more heterogeneous cohorts are required to confirm the reproducibility metrics reported here.

Second, all examinations were performed on two 3 T scanners from the same vendor using identical head coils and a fully harmonized spin-echo EPI-MRE protocol. The reported ARD, CV, RDC, and ICC values therefore characterize reproducibility under highly controlled, vendor-specific conditions. Furthermore, the acquisition order at *sites A* and *B* was kept consistent, and variations in this order were not assessed. The presented results may not directly extend to other field strengths, vibration driver concepts, sequence implementations, or inversion algorithms. Differences in reconstruction pipelines may introduce systematic offsets that equal or exceed the cross-site differences observed in this study. Previous work by Svensson *et al*. (2021) and Herthum *et al*. (2022) demonstrated that the choice of reconstruction method can substantially influence measured brain stiffness values as well as derived reproducibility metrics [24, 27].

Third, the study employed a specific multifrequency spin-echo MRE sequence with a defined set of vibration frequencies and wave-sampling parameters. Brain viscoelastic properties depend on vibration frequency, and modifications of the frequency spectrum or protocol design may alter both absolute mechanical property values and their cross-site variability [18, 21]. Consequently, the reproducibility metrics reported here apply only to the acquisition conditions used in this study.

Fourth, the analysis relied on a fixed set of atlas-based regions of interest. Small anatomical structures with limited voxel counts, complex microstructural organization, or proximity to the skull base are more susceptible to partial-volume effects and residual registration errors. These factors may contribute to the elevated variability observed in subcortical gray matter.

Finally, it is important to note that long-term stability of SWS and PR across days, scanner maintenance cycles, or hardware changes was outside the scope of this work, and was therefore not evaluated. Future studies may include larger and more heterogeneous cohorts, repeated measurements over extended intervals, additional scanner vendors and field strengths, and patient populations to establish robust and universally applicable benchmarks for brain MRE reproducibility.

## 6 Conclusion

This cross-site study demonstrates that brain multifrequency MRE provides robust and reproducible measurements of brain viscoelasticity across two 3 T MRI platforms when harmonized acquisition and processing protocols are applied. Overall, SWS and PR showed very good consistency across major brain tissue compartments, according to ARD, RDC, and CV metrics, and moderate-to-good agreement according to ICC, which is more sensitive to between-subject variability. White matter and cortical gray matter exhibited particularly stable measurements, whereas subcortical gray matter showed higher variability. Nevertheless, observed variability remained well below previously reported disease-related effect sizes for all compartments [11, 12, 6], which supports the application of brain MRE in multi-site research settings. Overall, the presented results indicate that brain multifrequency MRE enables reliable cross-site assessment of brain mechanical properties and provide benchmark reproducibility metrics that may facilitate future multi-center studies and clinical applications of brain MRE.

## Data Availability

All data produced in the present study are available upon reasonable request to the authors

## 7 Acknowledgements

This work was funded by the German Research Foundation (DFG) project 460333672 CRC1540 EBM. The authors would like to express their gratitude to all participants who volunteered for this study.

